# Analysis of spliceosome-related coding and noncoding genes and pseudogenes reveals novel candidates

**DOI:** 10.64898/2026.08.06.26358951

**Authors:** Olfa Messaoud, Stephanie DiTroia, Rahaf Tarawneh, Daniel Marten, Emily O’Heir, Melanie O’Leary, Lynn Pais, Vijay Ganesh, Moriel Singer-Berk, Broad CMG and GREGoR consortium collaborators, Monica Wojcik, Kaitlin E. Samocha, Heidi L. Rehm, Christina Austin-Tse, Anne O’Donnell-Luria

**Affiliations:** Center for Mendelian Genomics, Program in Medical and Population Genetics, Broad Institute of MIT and Harvard, Cambridge, Massachusetts, USA; Harvard Medical School, Boston, Massachusetts, USA; Division of Genetics and Genomics, Boston Children’s Hospital, Boston, Massachusetts, USA; Center for Genomic Medicine, Massachusetts General Hospital, Boston, Massachusetts, USA; Biomedical Genomics and Oncogenetics Laboratory, Institut Pasteur de Tunis, Tunisia; Harvard T.H. Chan School of Public Health, Massachusetts, USA; Department of Neurology, Brigham and Women’s Hospital, Boston, Massachusetts, USA; Division of Newborn Medicine, Boston Children’s Hospital, Boston, Massachusetts, USA

**Keywords:** Spliceosome genes, snRNAs, pseudogenes, neurodevelopmental disorder, retinal disorders

## Abstract

Splicing is a complex molecular mechanism in eukaryotic cells essential to gene expression and regulation, involving more than 300 protein-coding genes (PCGs) and 43 small nuclear RNA (snRNA) genes. However, fewer than 30 gene-disease relationships have been described as spliceosomopathies to date. This discrepancy suggests the splicing machinery as an underexplored area for human disease gene discovery.

For snRNA currently classified as pseudogenes, we prioritized candidates with similar epigenomic, genomic, and hypermutability features as functional snRNA genes. Population-variant-depletion analysis was performed to identify regions under negative selection. We analyzed rare variants in PCGs and snRNA genes and prioritized snRNA pseudogenes across a large heterogeneous rare disease cohort.

There was high concordance for prioritizing genes annotated as pseudogenes by the variant-depleted region analysis (9) and by random forest models of hypermutation, genomic and epigenomic features (6). We identified 26 variants of interest across six PCGs with established gene-disease relationships (GDRs) and 14 genes not yet disease-associated, including one pseudogene across 30 individuals. For snRNAs genes, we identified 49 variants of interest located in seven genes with established GDR and 11 genes not yet disease-associated, including two pseudogenes across 80 individuals.

This study highlights the importance of splicing-related PCG and snRNA in the genetic etiology of rare diseases. By leveraging specialized approaches for prioritizing pseudogenes, combined with the PCG and snRNA analysis, the genes and variants expand the variant pathogenicity spectrum of spliceosomopathies and suggest variants for follow-up case series and future functional validation.

## Introduction

Splicing is the processing of precursor messenger RNA (pre-mRNA) to mature messenger RNA (mRNA) and represents a critical molecular step in gene expression and regulation within eukaryotic cells. It is a complex molecular process involving proteins and RNA that form the ribonucleoproteins of the major and minor spliceosome, the main component of the splicing machinery ^1, 2^.

Several splicing-related small nuclear ribonucleoproteins (snRNP) gene-disease relationships (GDRs) have been described and termed spliceosomopathies ^3^. These include mainly retinal ^4^, neurodegenerative and neurodevelopmental syndromes ^5^, and craniofacial disorders, highlighting the importance of efficient and accurate splicing in development and cellular function ^6^.

Recently, there have been several reports of small noncoding RNA (snRNA) GDRs discovered, including several genes associated with distinct monogenic conditions with dominant and recessive inheritance modes. These are mainly associated with neurodevelopmental disorders (NDD) or retinal disease. One of these recent GDR discoveries led to the reclassification of a pseudogene as a gene —*RNU2-2P,* now renamed *RNU2-2* ^7^, highlighting the need to further evaluate snRNA pseudogenes.

Studies of major spliceosome snRNA components that are involved in splicing U2-type introns, which represent more than 99% of introns ^8^, have linked pathogenic variants in *RNU4-2*, *RNU5B-1*, and *RNU2-2* to several NDDs, including ReNU syndrome (MIM 620851), neurodevelopmental disorder with seizures and joint laxity (MIM 621302), and RNU2-2 developmental and epileptic encephalopathy 119 or ReNU2 syndrome (MIM 621304), respectively ^9, 10,11^. In addition, *RNU5A-1* is a candidate gene for an NDD ^10^. Recently, pathogenic variants in U4/U6 snRNA gene family have been associated with retinal degeneration (RD) ^12^.

Similarly, for the minor spliceosome, which is involved in splicing U12-type introns, representing ∼800 introns (<1%), two core snRNA genes are well established as disease-associated. *RNU4ATAC* is responsible for a group of diseases known as RNU4atac-opathy (MIM 226960, 210710, 616651) ^13,14^, and *RNU12* is associated with autosomal recessive spinocerebellar 33 ataxia (MIM 620208) ^15^ and CDAGS syndrome (MIM 603116) ^16^. *RNU6ATAC* has recently been associated with a neurodevelopmental disorder ^17^ with both *RNU4ATAC* and *RNU6ATAC* also associated with a syndromic monogenic autoimmune diabetes ^18^. While many diseases are associated with protein coding genes (PCGs) of the major spliceosome ^3^, only one minor spliceosome PCG is associated with disease (*RNPC3,* pituitary hormone deficiency, MIM 618160) ^19^.

Given the large number of PCGs and snRNAs involved in splicing, there is potential to discover new GDRs and splicing-related mechanisms. Here, we aim to expand prior approaches to evaluate variation in splicing components across research participants with unsolved rare disease sequenced through the Broad Center for Mendelian Genomics ^20^ and GREGoR (Genomics Research to Elucidate the Genetics of Rare Diseases) consortium ^21^. We evaluate noncoding genes that encode components of the major and minor spliceosome, the PCGs that interact with snRNA in splicing, and pseudogenes of these two gene classes ^22^. The main goal is to expand our knowledge of the splicing-related diseases by identifying additional diagnoses and candidates for further evaluation.

## Subjects and Methods

### Cohort

We collected rare disease data from the GREGoR consortium, the Broad Center for Mendelian Genomics, and other studies, which encompassed around 23,000 families (∼25,000 with exomes and ∼10,000 with genomes) originating from different geographical locations. These data were analyzed using the seqr analysis platform ^23^, and included participants affected with various rare diseases. These were predominantly neurodevelopmental and neuromuscular disorders but also included multiple congenital malformations and organ-based disorders such as bone marrow failure, retinal, vascular and renal disorders. This study was approved by the Massachusetts General Brigham IRB (protocols #2016P001422 and #2013P001477). All participants (patients or legal guardians) gave written informed consent via a local human subjects research protocol before data and sample collection.

### Genomic analysis

We focused our analyses on PCGs, snRNA noncoding genes, and pseudogenes for both gene sets that are putatively associated with splicing.

#### **-** Protein-Coding Genes and Pseudogenes

To identify PCGs involved in splicing, we interrogated the Reactome Pathway Database (https://reactome.org) with the keyword “splicing.” From a total of 716 proteins

involved in the metabolism of RNA, we retained 287 proteins that play a role in Processing of Capped Intron-Containing Pre-mRNA, among which 212 are involved in mRNA splicing. We included genes of the second level of this hierarchy for more comprehensive assessment. Among the set of 287 proteins, nine corresponded to isoform pairs or trios, encoded by the same gene (two isoforms for *CPSF6*, *SEH1L*, and *NUP58* and three isoforms for *NUP98*). Therefore, the total number of targeted genes was reduced to 282 genes. We added 129 pseudogenes related to these PCGs from HGNC ^24^. Of these, 38 genes were not considered a valid entry in seqr and were excluded from the search. The gene list was annotated for loss-of-function constraint metrics from the Genome Aggregation Database (gnomAD) v4.1.0. We excluded 21 genes without gnomAD constraint scores from the 282 PCGs, leaving 261 genes for analysis (Figure 1). We evaluated single nucleotide, insertion-deletions, or structural variants (SNV, indel, and SV, respectively) in the rare disease dataset. Fifty percent (131) of the PCGs had evidence of loss-of-function constraint (i.e., haploinsufficiency) indicated by pLI >0.9 (compared to ∼16% of all genes with high pLI). We evaluated all 261 PCGs and 91 pseudogenes for variants under *de novo*/dominant and recessive inheritance modes.

**Figure 1:**
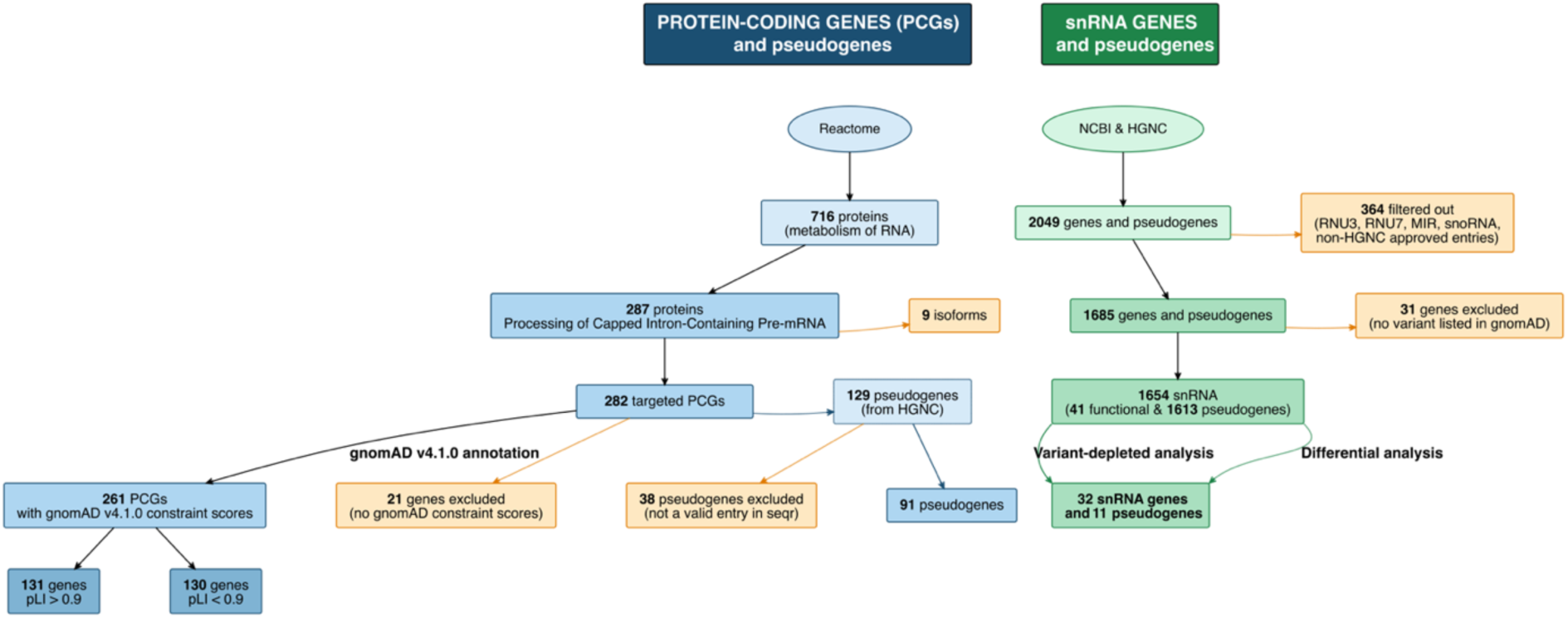
Flowchart illustrating the gene prioritization pipeline for protein-coding genes and snRNAs genes and pseudogenes. PCG and snRNA gene and pseudogene pipelines are shown in blue and green, respectively. Excluded genes are highlighted in orange. Circles represent web sources for gene collection and number of retained genes and pseudogenes at each step are shown in rectangles.

#### **-** snRNA genes and pseudogenes and the depletion strategy

The list and characteristics of snRNA genes and pseudogenes were downloaded from NCBI and checked for completeness by interrogating HGNC. From a starting list encompassing 2049 genes and pseudogenes, *RNU3*, *RNU7*, *MIR*, small nucleolar genes, and genes without a HGNC-approved status were excluded with1685 genes and pseudogenes remaining. At this step, 31 genes that had no variant listed in gnomAD were also excluded due to concerns for sequencing quality with short reads, yielding 1654 snRNA genes as our final gene dataset (41 are annotated as functional genes and 1613 as pseudogenes) (Figure 1). Data related to snRNA coordinates were retrieved from NCBI. These span the transcribed region from the Transcription Start Site (TSS) to the Transcription Termination Site (TTS). Since these genes do not contain introns, genomic locations from TSS to TTS correspond to RNA positions. Information on strand orientation was extracted from Ensembl (EnsDb.Hsapiens.v86) and biomaRt as well as manual curation for 11 pseudogenes.

As constraint data are not available for snRNAs, we identified regions of variant depletion in the general population using gnomAD v.4.1.0 genomes. Data related to allele frequency of 50,833 variants, encompassing 76,156 genomes, served to perform variant-depletion analysis. We converted these variants to their n. coordinates and discarded variants outside of the TSS-TTS boundaries, leaving 46,317 variants. These were then used to determine variant-depleted regions. We conducted a systematic grid search to identify optimal parameters for variant-depletion analysis. We tested seven window sizes (10, 12, 14, 16, 18, 20, and 25bp) across six depletion thresholds (0.10, 0.12, 0.14, 0.16, 0.18, 0.20), making a total of 42 combinations to classify windows as depleted vs non-depleted. For each combination, we applied sliding windows with a 1bp step across all genes, then calculated the distance of this window to the expected value (the SNV mean for each gene) following the formula: distance to expected = (observed SNVs / (window size × 3)) - (expected rate × window size / (window size × 3)). We multiplied by 3 to account for the three possible single-point mutations at each nucleotide. Depleted regions were defined as those where the distance is less than the evaluated threshold. To evaluate the statistical significance of the defined depleted regions, we compared minimum distance values between depleted and non-depleted genes using Wilcoxon test with the effect size determined using Cohen’s d.

To determine the optimal combination, we opted for the threshold that overlapped with the inflection point (0.14) and the window size that resulted in the highest combined score (10bp) that we defined based on statistical parameters (combined score = the significance weight + effect size + stability + reasonable detection rate). Genes harboring at least one depleted window were prioritized for variant search. Regions depleted in variants were identified by their physical coordinates. These regions are potential candidate loci for dominant pathogenic variants.

Finally, the presence of any rare variant in these regions of interest among affected individuals was assessed for snRNA genes and pseudogenes within depleted regions. To ensure a comprehensive coverage of genes with variant-depleted regions, we extended the analysis to the full gene sequence for both dominant and recessive searches. For *de novo*/dominant variant filtering, we mainly retained ClinVar pathogenic variants and variants annotated as noncoding transcript exon variants with a gnomAD allele frequency ≤0.1%, seqr allele count ≤20, genotype quality ≥20, and allele balance ≥0.2. For recessive variants, we used the same variant annotation criteria and quality metrics but applied a gnomAD allele frequency ≤0.5% and seqr allele count ≤200.

#### **-** Differential analysis of genes vs pseudogenes

Based on the recent reclassification of *RNU2-2* from a pseudogene to a functional gene, we wanted to assess pseudogenes more broadly for evidence of functionality. We performed a random forest (RF) analysis based on differential characteristics between genes and pseudogenes. These characteristics are classified into three categories that are related to genomic features, epigenomic features, and hypermutability. Data related to 5’ and 3’ genomic annotations were extracted from UCSC (https://genome.ucsc.edu/), epigenomic data from Encode (https://www.encodeproject.org/), and variant-related data from gnomAD v4.1.0 (Supplementary Tables 1-3).

For genomic features, we gathered information related to gc_promoter and cpg_density for promoter/regulatory regions (2000bp upstream of the gene) and data on polya_canonical, gc_3prime, and u_richness for 3′-processing signals (500 bp downstream of the gene).

For epigenetic features, values representing the normalized peak intensity of H3K4me3 histone marks for promoters, H3K27ac and H3K4me1 histone marks for active enhancers were extracted. We selected results of Histone CHIP-seq by filtering specifically from neuronal tissues (e.g., brain, dorsolateral prefrontal cortex, and cerebellum tissues). Most of the datasets (125/128) were from adults, except for three, which were derived from embryonic tissues.

For hypermutability, we first checked whether snRNA functional genes have statistically significant differences in the number of variants per gene and the normalized-to-gene-size number of variants compared to pseudogenes (Wilcoxon rank-sum test). Then, we assessed other distinctive characteristics including the number of variants per gene and their frequencies according to the high-quality allele counts from gnomAD v4.1.0, the transition vs transversion proportion, and the number of singletons. We also evaluated the presence of a specific variant positional distribution along the transcripts (Kolmogorov-Smirnov test).

To identify pseudogenes with gene-like characteristics, we performed Random Forest (RF) analyses on the following three feature categories: hypermutation features (variant count, variants per kilobase, proportion of singletons, Transitions/Transversions ratio, mean allele frequency), genomic features (CpG density, promoter GC content, poly-A signal, 3’ GC content, uridine richness), and epigenomic features (H3K4me3, H3K27ac, H3K4me1). For each category, an RF model was trained using 1000 trees and a balanced sampling to guarantee a stable prediction. Performance was evaluated by repeated 10 × 5-fold cross-validation, and pseudogenes with a P(Gene) > 0.5 (as a default threshold) were classified as gene-like within the tested domain. Mean decrease in accuracy was measured for each feature, with higher values attributed to important features. This analysis was performed independently for each feature category, and pseudogenes meeting criteria across all three categories were prioritized as high-confidence candidates, while pseudogenes meeting criteria in two or one category were classified as medium- and low-confidence candidates, respectively. To validate the results obtained from separate features, we trained an additional model combining all features together.

#### **-** Comparison of prioritized pseudogenes

For pseudogenes prioritized as gene-like candidates based on either the variant-depletion analysis or the random forest-based differential analysis, we assessed the concordance between the two approaches via enrichment and correlation analyses. Variant-depletion analysis prioritizes pseudogenes that contain a depleted region; these are quantified by the number of depleted regions, total depleted base pairs, largest depleted region, mean region size, and strongest depletion signal. The differential approach classifies pseudogenes based on hypermutation, genomic, and epigenomic features using three levels of confidence (high, medium, and low) that are proportional to the number of features being met. First, we tested whether pseudogenes retained by the depletion analysis were enriched for differential classifications by comparing those retained by at least one feature with those not selected at all using Fisher’s exact test. Second, for pseudogenes selected by the two approaches, we examined their distribution across the different confidence levels assigned by the differential analysis. Finally, for pseudogenes prioritized by the first approach, we assessed correlations between the five quantitative depletion parameters (cited above) and the evidence levels of the differential approach. The latter were treated as an ordinal variable (none=0, low=1, medium=2, high=3) using Spearman’s rank correlation. To visualize the overlap between the three features of the differential approach (hypermutation, genomic, and epigenomic) and the depletion approach, a proportional Venn diagram was constructed using the euler R package ^25^.

All statistical analyses were performed in R (version 4.5.1) using base statistical functions and the dplyr package for data manipulation. Statistical significance was set at α = 0.05.

#### **-** Variant analysis

Variant filtering and review were performed using the seqr platform. For *de novo*/dominant variant filtering in PCGs, we retained predicted loss-of-function (pLoF) (e.g., nonsense, frameshift, essential splice site), missense and inframe indel variants segregating in trios or among two affected family members that met the following criteria: a gnomAD v4.1 allele frequency ≤0.01%, seqr internal allele count ≤10, deleterious computational predictor scores (CADD ≥20, SpliceAl ≥0.2, or REVEL ≥0.75), and passing quality metrics (genotype quality ≥30, and allele balance ≥0.2). For recessive variants, we applied the same criteria as for *de novo* inheritance, except for using more relaxed gnomAD frequency and seqr allele count with a threshold of 1% and 3000, respectively, and a more stringent allele balance of 0.25. For SV, we retained only pLoF SVs that are absent from gnomAD, with a seqr allele count ≤3, and quality metrics (≥50 for WES SV Quality Score and ≥5 for WGS SV Quality Score). Phasing was assessed for bi-allelic variants when possible. For snRNA genes, in addition to diagnostic variants, we also retained variants that fulfill prioritization criteria without evidence of familial segregation. We were also interested in the idea of amorphic (complete loss of gene function) and hypomorphic (partial loss of gene function) compound heterozygous variants, as there is some suggestion in the field that these combinations may be causal of disease. As we do not have data on the functional impact of individual variants, we are using allele frequency as a proxy, where we considered variants that are absent from gnomAD in the homozygous state as amorphic and reported as rare but homozygotes present in gnomAD for the second allele as hypomorphic.

Variants were annotated and classified according to ACMG/AMP and ClinGen criteria ^26, 27^. Whenever RNAseq data were available, splice junction outliers (SJO), as a typical feature for some spliceosomopathies, were evaluated using FRASER2 in addition to checking for expression outliers using OUTRIDER ^28^.

To collect additional cases to strengthen evidence for a gene-disease association, for candidate genes not yet associated with disease, at least one case per gene with their relevant HPO terms and candidate variants, was shared with the scientific community via Matchmaker Exchange Network ^29^ using the seqr node.

## Results

The set of PCGs, snRNA genes, and pseudogenes used for variant analysis were prioritized using the pipeline illustrated by Figure 1.

### - Protein-coding genes (PCG) analysis

Variant assessment across 261 genes and 91 pseudogenes revealed 26 candidate variants among 30 individuals from 26 families with rare diseases, mostly NDD, but also other phenotypes such as neuromuscular diseases and microcephaly (Supplementary Table 4). Eight missense, two nonsense, one deletion, one frameshift, and one intronic *de novo* variants in 11 genes (10 of which were loss-of-function constrained) were identified in 12 patients (one patient had two missense variants in two genes: *SF3A2* and *SNW1*). In addition, we identified a dominantly inherited missense variant in *CWF19L2* gene shared by a mother and her daughter, both with an NDD phenotype. We also identified a nonsense variant in *CWC25* gene shared by 3 sibs with NDD. Two variants per gene were identified in *SRRT* and *SRRM2* genes.

Five homozygous variants and one compound heterozygous variant (four missense, one synonymous, one intronic and one nonsense) in four genes among six patients were also identified. No recurrent variant was observed among biallelic variants. However, we noted allele heterogeneity for *NUP210* and *CPSF1* gene, each with two variants.

For SVs, three *de novo* deletions spanning different genes were identified in three individuals. For pseudogenes, recessive and SV searches yielded zero variants while the dominant search identified 44 variants. Two SNV were prioritized, both in *BUD13P1,* in three patients with muscular dystrophy.

### - snRNA gene and pseudogene analysis

#### · Variant-depletion results

A total of 1654 snRNA genes and pseudogenes that form the major and the minor spliceosome were evaluated, with 41 being annotated as genes and 1613 as pseudogenes. Grid search and optimal combination analysis yielded a threshold for depletion of 0.14 (i.e., a deviation from the mean of at least 14%) and a window size of 10 (Figure 2-a; Supplementary Figure 1). Variant-depleted region analysis using these parameters showed the presence of 71 regions located within 41 genes and pseudogenes. Among these, 11 were previously described as disease-associated genes (DAG) with an established or suggested relationship (*RNU2-2, RNU4-2, RNU5A-1, RNU5B-1, RNU6-1, RNU6-2, RNU6-8, RNU6-9, RNU12, RNU4ATAC, RNU6ATAC)*, 21 not yet disease-associated (*RNU1-1, RNU1-2, RNU4-1, RNU5D-1, RNU5E-1, RNU5F-1, RNU11, RNVU1-2A, RNVU1-3, RNVU1-6, RNVU1-7, RNVU1-8, RNVU1-14, RNVU1-15, RNVU1-18, RNVU1-19, RNVU1-20, RNVU1-21, RNVU1-22, RNVU1-24, RNVU1-34),* and 9 pseudogenes (*RNU1-108P, RNU1-154P, RNU2-63P, RNU5A-8P, RNU5E-4P, RNU5E-6P, RNU6-1151P, RNU6-328P, RNU6-457P*) (Figure 2-b). The number of depleted regions per gene ranged from one to three with the total depletion within a gene showing a maximum of 54 bp, and the largest individual depletion region was 41 bp. Their distribution throughout each gene shows focal depletions for some and global depletions for others (Figure 2-c; Supplementary Table 5; Supplementary Figure 2).

**Figure 2:**
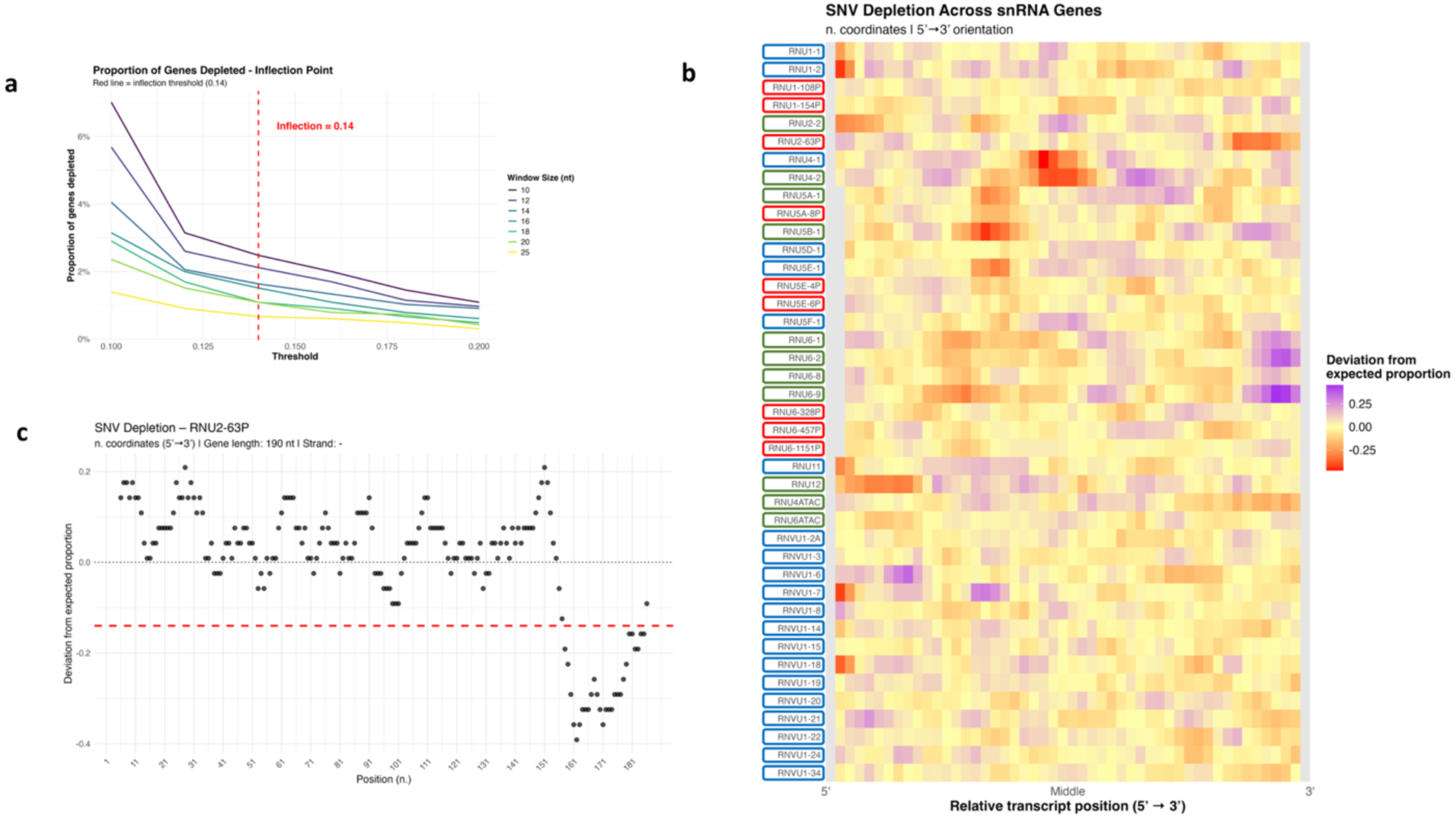
Variant-depletion strategy optimization and implementation. a) Identification of the optimal depletion threshold by assessing the proportion of genes with depletion across different thresholds and for variable window sizes. b) Depletion heatmap at the defined optimal parameters (window size = 10bp and depletion threshold = 0.14) showing 71 variant-depleted regions across 41 genes (red = depleted; yellow = expected; purple = enriched). Genes with established gene-disease relationship (green), genes not yet disease associated (blue), and pseudogenes (red). c) Normalized SNV proportion showing a 37 bp depleted region close to the 3’ end of the *RNU2-63P* pseudogene.

#### · Differential analysis between genes and pseudogenes

There is evidence that some snRNA pseudogenes may be misannotated and may be functional genes. For example, *RNU2-2P* was recently renamed as *RNU2-2* and both a dominant and recessive GDR has been described. To prioritize other pseudogenes that may be misannotated for further analysis, we conducted a differential analysis based on genomic features, epigenomic signatures and gene hypermutability characteristics. We started with a comparison between genes and pseudogenes that showed that snRNA genes have statistically significantly higher number of variants than pseudogenes (Wilcoxon P = <2e-16), a feature that persisted even after normalization to gene length (Wilcoxon P = <2e-16) and might result from the high transcription rate of active snRNA genes, especially those transcribed by polymerase III ^30^. This significant difference was also observed for the other evaluated features: transition vs transversion proportion (Chi-square P = <2e-16), the number of singletons in gnomAD v4.1 (Wilcoxon P = 8.97e-14), and allele frequency (Chi-square P = <2e-16). In addition, the comparison of the variant position distribution showed a significant difference in the distribution uniformity marked by a pronounced depletion in the 5’ and 3’ ends of the transcripts (KS P = 0.0392) (Supplementary Figure 3).

Discriminative performance evaluation showed high performance for the three RF models to distinguish between snRNA genes and pseudogenes with AUC = 0.999 (hypermutation), 0.996 (genomic), and 0.995 (epigenomic) (Figure 3-a). Using P(Gene) > 0.5 as a cutoff yielded 28 gene-like pseudogenes for hypermutation, 85 for genomic, and 80 for epigenomic features (Supplementary Table 6). When converging the three RF models, we retained six high-confidence gene-like pseudogenes (*RNU5E-4P, RNU2-63P, RNU5E-6P, RNU1-5P, RNU1-108P,* and *RNU1-6P*) that we prioritized for variant analysis. Twenty-two additional pseudogenes were classified with medium-confidence and 131 pseudogenes had low-confidence. Results of the combined RF model (AUC = 0.999) confirmed findings of the three separate models, where the five pseudogenes with the highest combined scores showed high-confidence based on the convergent model (Figure 3-b; Supplementary Table 6).

**Figure 3:**
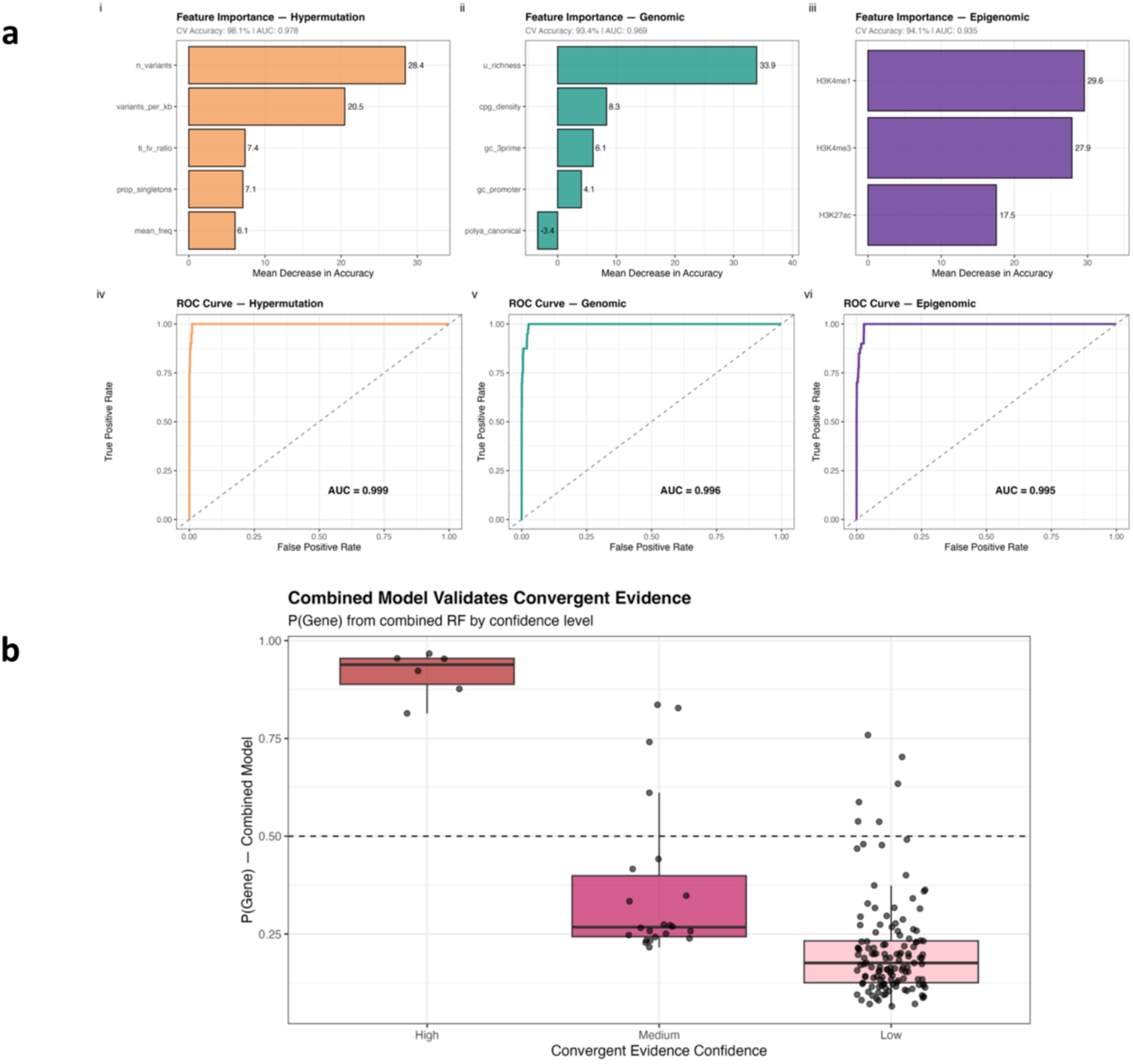
Differential analysis between genes and pseudogenes. a) Top panel shows the variables included in each feature and their importance illustrated by mean decrease in accuracy. Bottom panels show ROC curves and high AUC values (all ≥ 0.995) indicating the ability of all features to significantly differentiate genes from pseudogenes. b) Combined model validating the convergent evidence results: dots represent combined scores for pseudogenes classified according to their convergent evidence.

#### · Comparison of prioritized pseudogenes

To assess concordance between the two pseudogene prioritization approaches, we performed enrichment and correlation analyses. Results showed that pseudogenes retained by the variant-depletion approach were enriched for pseudogenes classified as gene-like by the differential approach (Fisher’s P = 6.6e-8), with an overlap of 89% (8/9). Among these, the variant-depletion approach preferentially prioritized pseudogenes with high evidence level, with a rate of 44% (Supplementary Figure 4-a). However, no significant correlation between depletion parameters and evidence levels was noted. The proportional Venn diagram showed four pseudogenes being selected by all features of the differential approach and the depletion strategy (*RNU5E-6P, RNU5E-4P, RNU2-63P,* and *RNU1-108P*) and the greatest overlap was for the hypermutation feature (Hypermutation ∩ Depletion =8 vs Epigenomic ∩ Depletion = 5 and Genomic ∩ Depletion =4) (Supplementary Figure 4-b).

#### · Analysis of prioritized snRNA genes and pseudogenes

Variant analysis of both 32 snRNA genes and 11 prioritized pseudogenes, retained based on variant-depletion analysis or as gene-like, retained based on the differential analysis (except for *RNVU1-20* which was not recognized by seqr), allowed the identification of 49 unique variants (seven *de novo*, two dominant, 16 homozygous, 19 compound heterozygous, three amorphic and hypomorphic compound heterozygous, and two unknown) in 17 genes (7 genes with established GDRs and 11 candidate genes including two prioritized pseudogenes; *RNU4-2* gene is counted in both classes) in 80 patients (Supplementary Table 7; Supplementary Figure 5).

##### 1- snRNA genes with established GDR

We identified 22 deleterious variants (six *de novo*, five homozygous, and 11 compound heterozygous) with a recurrent variant (*RNU4-2*: n.64_65insT identified in 15 patients) which allowed us to establish a genetic diagnosis for 43 patients, many of whom have been previously published in the original or follow-up GDR discovery manuscripts. These pathogenic variants are located in five genes: *RNU2-2*, *RNU4-2*, *RNU5B-1*, *RNU4ATAC,* and *RNU6ATAC*. All these were previously described to be associated with NDD, thus confirming the reliability of our analysis pipeline. Furthermore, we identified 11 candidate deleterious variants in genes with established GDR (*RNU2-2*, *RNU4-2*, *RNU5A-1*, *RNU4ATAC,* and *RNU12*) in 16 patients. These were six inherited homozygous, three inherited compound heterozygous, one inherited amorphic and hypomorphic compound heterozygous, and one heterozygous of unknown inheritance. Functional effect was confirmed for some variants based on transcriptomic analysis showing high number of splice junction outliers (Supplementary table 7; Supplementary figure 6) and/or supporting segregation data ^10,11,31–34^.

##### 2- snRNA genes with candidate GDR

From the list of candidate variants, n.18_19insA in *RNU4-2* was identified in a multiplex family where the father presented with isolated RD and his three children presented with NDD and RD. This variant, mapped to the three-way junction of the complex RNU4/RNU6, has been previously described in patients with RD alone.^9^ While no additional variants of interest were identified in the children to account for the NDD, it remains unclear if we are seeing a phenotype expansion with variable expressivity or if this is a partial solve with the *RNU4-2* variant accounting for the RD.

In addition to *de novo* variants in *RNU5B-1*, we identified biallelic variants in *RNU5A-1* previously suggested to be associated with monoallelic NDD. We also retained biallelic variants in another RNU5 gene, *RNU5E-1*, and a pseudogene, *RNU5E-6P*. Furthermore, we identified four biallelic variants in *RNU5F-1* gene in patients with NDD, among them two with coloboma. This gene revealed an enrichment in unsolved cases for both *de novo* and biallelic variants. All candidate biallelic variants in *RNU5F-1* gene were clustered in the stem I region, except one allele which was located in the U5 5’ loop I, part of the 5’ exon recognition site.

We also retained one candidate variant in *RNU2-63P*. This pseudogene, despite being among the most hypermutable splicing-related snRNAs, contains the second largest variant-depleted region (after *RNU4-2)* encompassing 37 bp, which is located in the 3’ end (Figure 2-c). It is also the sixth strongest depletion (Supplementary Table 5), which reflects high constraint. Furthermore, it was reclassified as gene-like with high confidence by the differential analysis. *RNU2-63P* showed an enrichment in unsolved cases for both *de novo* and biallelic variants.

Additional cases and more careful evaluation of phenotypes will be needed to further evaluate these candidates.

## Discussion

Understudied molecular and cellular pathways represent opportunities worth exploring to unravel part of the underlying genetic etiology of ultra-rare and undiagnosed diseases. In this context, we conducted a comprehensive genetic analysis of spliceosome-related PCGs, noncoding genes, and their pseudogenes across a large, rare disease cohort. As for coding genes, prioritization of monoallelic variants was based on constraint measures, computational predictors, and variant segregation. Even in a cohort of this size, few patterns emerged through a number of candidate variants that were identified and combined with other large cohorts towards establishing novel GDRs and variant pathogenicity.

While ACMG/AMP and ClinGen standards exist for variant classification for PCGs, interpretation of variants in noncoding genes was challenging. Hence, adapted and combined strategies were leveraged. Specifically, we evaluated regional burden to identify areas depleted for variation in the human population and further extended the search to all gene sequences to uncover potential recessive inheritance.

In addition to snRNA genes, we prioritized a list of pseudogenes based on variant-depleted region analysis on the one hand and common hypermutation, genomic, and epigenomic features shared with genes on the other. Absence of correlation between the two pseudogene reclassification approaches indicated that variant-depletion strategy and differential analysis captured different characteristics and the two methods are complementary. This non-redundant behavior supported the use of gene-like pseudogenes prioritized by the two approaches as candidates for future evaluation with a focused attention on overlapping ones. For these, we hypothesize that some might be misclassified, but additional genetic and functional evidence is needed to confirm their disease association. Caution is warranted when dealing with non-neurological phenotypes since the epigenomic features we selected were designed for genes important in the nervous system and may miss a role in other organ systems.

Genetic analysis showed that the majority of snRNA variants segregate in a recessive pattern, which is consistent with the high frequency of biallelic variants described in other splicing-related snRNA genes responsible for neurodevelopmental disorders. However, it is important to note that the mutation rate in these genes appears to be high, resulting in a higher frequency of variation in snRNA genes than in PCGs. Our preliminary results related to the RNU5 family endorse this observation and suggest consideration of expansion of the inheritance mode from dominant to recessive for a candidate NDD-related gene (*RNU5A-1)*. This inheritance expansion has been recently described for *RNU4-2* and *RNU2-2* genes, both originally reported as associated with dominant NDD, but later found to also have recessive forms ^7,9,11,31–33,35^. Similarly, the identification of the variant n.18_19insA in *RNU4-2,* previously associated with RD, in a family with NDD and RD suggests either a missed dual diagnosis with a blended phenotype or a phenotype expansion/variable expressivity associated with this variant.

Considering the absence of a clear transcriptomic or epigenomic feature that characterizes RNU deficiency for most major spliceosome genes, and because computational predictors are biased towards the coding portion of the genome ^36^, we limited our analysis to variant clustering and localization. For candidate variants in *RNU5F-1* gene, the presence of at least one allele of the biallelic variants in the stem region, which is crucial to U4/U6.U5 tri-snRNP formation, suggests a common pathogenic disease mechanism and further supports GDR candidacy for this gene. This domain is not only involved in maintaining a stable RNU5 structure to allow U5 loop 1 positioning at the active site ^37^, but it is also part of a larger region that interacts with other snRNPs proteins to bring the RNU5 to the U4/U6.U5 tri-snRNP complex during the catalytic phase of splicing ^38^.

Conversely, evidence of pathogenicity was limited for most of the genes for which only one or two variants were identified. This is likely due to knowledge gaps in the field, resulting in few candidate variants per gene especially with the highly conservative filters applied. Indeed, the cutoffs used in this study are relatively stringent compared to those previously reported, such as the recent article by Leitão et al, in which they adopted AC<50 for *de novo* variants vs AC≤20 in our study. Furthermore, mapping issues for most of the investigated genes, especially those part of the major spliceosome, given their organization in tandem repeats or their high paralog similarity, complicate accurate read mapping and variant calling in short-read sequencing ^39^. This would lead to a lack of or incorrect (sequencing artifacts) information at certain loci, impacting comparison across all patients and genes. In addition, the diversity of phenotypes available is relatively limited for each condition, because they are all rare or ultra-rare conditions. This represented a major limitation for variant validation and phenotype-genotype correlation establishment, even with leveraging the Matchmaker Exchange Network and other collaborative networks.

Despite the limited findings for each gene, collectively snRNA deficiencies are responsible for a substantial number of NDD cases. Furthermore, the diversity of phenotypes seems to be wider than what is currently reported, especially after the recent discovery associating them with retinal degeneration ^12^ and syndromic monogenic autoimmune diabetes ^18^, highlighting their pleiotropic characteristics. Given that transcriptomic data showed that 1-2% of patients in our cohort have significant SJO scores (unpublished data), we anticipate that a transcriptomic-oriented analysis would unravel part of this system complexity.

The recent surge of studies associated with spliceosomes allowed the identification of new genes, new variants, new inheritance modes, and even pseudogene re-classification. There is little overlap in the candidate variants presented here and prior RNU publications (aside from the previously published cases from this cohort), which may be related to the hypervariability of RNU genes and consistent with significant allelic heterogeneity for these RNU-associated disorders that still needs to be further characterized (and differentiated from benign population variation).

In conclusion, this study highlights the importance of splicing-related protein-coding genes and noncoding snRNAs genes in rare diseases. It prioritizes a number of snRNA pseudogenes for more careful evaluation. Some of the snRNUs remain challenging to assess given high homology, and we anticipate that future studies using long-read sequencing and innovative analytical approaches will further demonstrate that noncoding RNA genes are major contributors to human health and disease.

## Data Availability

Genomic and phenotypic data from the Broad CMG are available via dbGaP accession numbers phs003047 (GREGoR) and phs001272 (CMG). Access is managed by a data access committee designated by dbGaP and is based on intended use of the requester and allowed use of the data submitter as defined by consent codes.

## Acknowledgements

We thank the families who participate in these research studies, the many clinicians and researchers involved in the research studies and participant recruitment, and former members of the Broad CMG.

## Authors’ contributions

O.M. analyzed data and drafted the paper. S.D., E.O., M.O., L.P., V.G., analyzed rare disease cases and/or evaluated variant selection. A.P. and S.A. provided project management. R.T. contributed to statistical analysis. D.M. extracted variant data from gnomAD. M.S.B and E.O. curated variant according to ACMG/AMP criteria. Broad CMG and GREGoR consortium collaborators recruited and reviewed cases for candidate variants. K.E.S. helped in the differential analysis design. H.R co-directs the Broad CMG and ensured funding. C.A-T. co-supervised the work. A.O-L. designed the project, co-directs the Broad CMG, ensured funding, provided critical reading of the article and supervised the work. All authors edited the manuscript and approved the final version.

## Declarations

### Ethics approval and consent to participate

This study was approved by the Massachusetts General Brigham IRB (protocols #2016P001422 and #2013P001477). All participants gave written informed consent via a local human subjects research protocol before data and sample collection.

### Consent for publication

All individuals included in this study participated in a research study and provided informed written consent.

### Data & Code Availability

Scripts used to analyze data and generate tables and figures are available as supplementary files.

## Broad CMG and GREGoR consortium collaborators

Amina Abubakar^1,2,3^, Abdulrahman Ahmed Aldeeri^4,5,6^, Katherine N Anderson^7^, Muhammad Ayaz^8,9^, Brenda J Barry^10,11,12^, Alan H Beggs^7,6,13^, Seth I Berger^14,15^, Jonathan A Bernstein^16^, Brian P Brooks^17^, Zandre Bruwer^18^, Kinga M Bujakowska^13,63,64^, Daniel G Calame^20,21,22^, Jessica X Chong^23,24^, Wendy K Chung^25,26^, Emmanuèle C Délot^27^, Kirsten A Donald^18,19^, Elizabeth C Engle^10,11,12,28^, Karen J Fieggen^18,19,29^, Lyndon Gallacher^30,31^, Casie A Genetti^7^, Richard A Gibbs^21^, Joseph G Gleeson^32,33^, Bin Guan^17^, Rita Horvath^34^, Robert B Hufnagel^17,35^, Muhammad Ilyas^8^, Patricia Kipkemoi^1,36^, Nicolai Kohlschmidt^37,38^, Hanns Lochmuller^39,40,41^, James R Lupski^21,22,42,43^, Alan S Ma^44,45^, Rodrigo Mendez^46^, Ganeshwaran H Mochida^5,7,47,48^, Stephen B Montgomery^49,50,51^, Jennifer E Neil^5,52^, Charles RJ Newton^1,2,3^, Kaisa T Oja^53,54^, Katrin Õunap^53,54^, Eric A Pierce^13,63,64^, Jennifer E Posey^55^, Andreas Roos^41,56,57,58^, Tony Roscioli^59^, Riccardo Sangermano^63,64^, Tipu Sultan^65^, Tiong Y Tan^30,31^, Ana Töpf^60^, Deniz Torun^66^, Ehsan Ullah^17^, Eric Vilain^27^, Christopher A Walsh^5,6,7,10,11,13,53,61^, Matthew T Wheeler^46^, Susan M White^30,31^, Changrui Xiao^62^, Elif Yilmaz Gulec^67,68^, Maha S Zaki^69^

Broad CMG and GREGoR consortium collaborators’ affiliations

1. Institute for Human Development, Aga Khan University, Nairobi, Kenya

2. Neuroscience Unit, KEMRI-Wellcome Trust, Kilifi, Kenya

3. Department of Psychiatry, Oxford University, Oxford, UK

4. Department of Medicine, College of Medicine, King Saud University, Riyadh, Saudi Arabia

5. Division of Genetics and Genomics, Boston Children’s Hospital, Boston, MA, USA

6. Harvard Medical School, Boston, MA, USA

7. The Manton Center for Orphan Disease Research, Division of Genetics and Genomics, Boston Children’s Hospital, Boston, MA, USA

8. Centre for Omic Sciences, Islamia College University, Peshawar, KP, Pakistan

9. Center of Biotechnology and Microbiology, University of Peshawar, Peshawar, KP, Pakistan

10. F.M. Kirby Neurobiology Center, Boston Children’s Hospital, Boston, MA, USA

11. Howard Hughes Medical Institute, Boston Children’s Hospital, Boston, MA, USA

12. Department of Neurology, Boston Children’s Hospital, Boston, MA, USA

13. Program in Medical and Population Genetics, Broad Institute of MIT and Harvard, Cambridge, Massachusetts, USA

14. Research and Development, Ambry Genetics, Aliso Viejo, CA, USA

15. Genetics and Metabolism, Children’s National Hospital, Washington, DC, USA

16. Department of Pediatrics, Division of Medical Genetics, Stanford University School of Medicine, Stanford, CA, USA

17. Ophthalmic Genetics and Visual Function Branch, National Eye Institute, National Institutes of Health, Bethesda, MD, USA

18. Department of Paediatrics & Child Health, Red Cross War Memorial Children’s Hospital, Cape Town, South Africa

19. Neuroscience Institute, University of Cape Town, Cape Town, South Africa

20. Section of Pediatric Neurology and Developmental Neurosciences, Department of Pediatrics, Baylor College of Medicine, Houston, TX, USA

21. Human Genome Sequencing Center, Baylor College of Medicine, Houston, TX, USA

22. Texas Children’s Hospital, Houston, TX, USA

23. Department of Pediatrics, Division of Genetic Medicine, University of Washington, Seattle, WA, USA

24. Brotman Baty Institute, Seattle, WA

25. Department of Pediatrics, Boston Children’s Hospital, Boston, MA, USA

26. Harvard Medical School, Boston, Massachusetts, USA

27. Institute for Clinical and Translational Science, University of California Irvine, Irvine, CA, USA

28. Departments of Neurology and Ophthalmology, Harvard Medical School, Boston, MA, USA

29. Department of Medicine Division of Human Genetics, University of Cape Town, Cape Town, South Africa

30. Murdoch Children’s Research Institute, Victorian Clinical Genetics Services, Melbourne, Victoria, Australia

31. Department of Paediatrics, The University of Melbourne, Melbourne, Victoria, Australia

32. Department of Neurosciences, University of California, San Diego, CA, USA

33. Rady Children’s Institute for Genomic Medicine, San Diego, CA, USA

34. Department of Clinical Neuroscience, Cambridge University, Cambridge, UK

35. Center for Integrated Health Care Research, Kaiser Permanente Hawai’i, Honolulu, HI, USA

36. Center for Genomic Medicine, Massachusetts General Hospital, Boston, MA, USA

37. Institute of Clinical Genetics and Tumour Genetics Bonn, Bonn, Germany

38. National Center of Genetics (NCG), Dudelange, Luxembourg

39. Department of Medicine, Division of Neurology, The Ottawa Hospital, Ottawa, Ontario, Canada

40. CHEO Research Institute, Ottawa, Ontario, Canada

41. Brain and Mind Research Institute, University of Ottawa, Ottawa, Ontario, Canada

42. Department of Molecular and Human Genetics, Baylor College of Medicine, Houston, TX, USA

43. Department of Pediatrics, Baylor College of Medicine, Houston, TX, USA

44. Department of Clinical Genetics, Sydney Children’s Hospitals Network - Westmead, Sydney, NSW, Australia

45. Specialty of Genomic Medicine, University of Sydney, Sydney, NSW, Australia

46. Department of Medicine, Division of Cardiovascular Medicine, Stanford University, Stanford, CA, USA

47. Pediatric Neurology Unit, Department of Neurology, Massachusetts General Hospital, Boston, MA, USA

48. Department of Pediatrics, Harvard Medical School, Boston, MA, USA

49. Department of Pathology, Stanford University, Stanford, CA, USA

50. Department of Genetics, Stanford University, Stanford, CA, USA

51. Department of Biomedical Data Science, Stanford University, Stanford, CA, USA

52. Howard Hughes Medical Institute, Boston Children’s Hospital, Boston, MA, USA

53. Institute of Clinical Medicine, University of Tartu, Tartu, Estonia

54. Genetics and Personalized Medicine Clinic, Tartu University Hospital, Tartu, Estonia

55. Department of Molecular and Human Genetics, Baylor College of Medicine, Houston, TX, USA

56. Center for Translational Neuro- and Behavioral Sciences, University Duisburg-Essen, Department of Neuropediatrics and Neuromuscular Centre for Children and Adolescents, Essen, Germany

57. Department of Neurology, Heinrich Heine University Düsseldorf, Düsseldorf, Germany

58. Department of Medicine, Division of Neurology, Children’s Hospital of Eastern Ontario Research Institute, Ottawa, Ontario, Canada

59. Randwick Genomics Prince of Wales Hospital, NSW Health Pathology, Neuroscience Research Australia (NeuRA), University of New South Wales, Sydney, NSW, Australia

60. John Walton Muscular Dystrophy Research Centre, Newcastle University and Newcastle Hospitals NHS Foundation Trust, Newcastle Upon Tyne, UK

61. Pediatrics and Neurology, Harvard Medical School, Boston, MA, USA

62. Neurology, University of California Irvine, Irvine, CA, USA

63. Department of Ophthalmology, Ocular Genomics Institute, Massachusetts Eye and Ear, Boston, MA, USA

64. Department of Ophthalmology, Harvard Medical School, Boston, MA, USA

65. Department of Paediatric Neurology, The Children’s Hospital and the University of Child Health Sciences, Lahore, Pakistan

66. Department of Medical Genetics, University of Health Sciences, Gulhane Faculty of Medicine, Ankara, Turkey

67. Department of Medical Genetics, Istanbul Medeniyet University, School of Medicine, Istanbul, Turkiye

68. Medical Genetics Clinic, Goztepe Prof Dr Suleyman Yalcin City Hospital, Istanbul, Turkiye

69. Clinical Genetics Department, Human Genetics and Genome Research Institute, National Research Centre, Cairo, Egypt

## Broad CMG and GREGoR consortium collaborators’ funding

A.A. NeuroDev is supported by the Stanley Center for Psychiatric Research at the Broad Institute. NeuroDev was also supported by a grant from the Simons Foundation or the Simons Foundation International (599648). Research reported in this publication was supported by the: National Institute Of Mental Health of the National Institutes of Health under Award Number U01MH119689; Eunice Kennedy Shriver National Institute Of Child Health & Human Development of the National Institutes of Health under Award Number R01HD102975; National Human Genome Research Institute of the National Institutes of Health under Award Number R01HG012781. The content is solely the responsibility of the authors and does not necessarily represent the official views of the National Institutes of Health. Sequencing was provided by the Broad Institute of MIT and Harvard Center for Mendelian Genomics (Broad CMG) and was funded by the National Human Genome Research Institute; the National Eye Institute; the National Heart, Lung, and Blood Institute grant UM1HG008900; and in part by National Human Genome Research Institute grant R01HG009141. A.H.B. Boston Children’s Hospital IDDRC Molecular Genetics Core Facility funded by P50HD105351 from the NICHD, and the Boston Children’s Hospital Children’s Rare Disease Collaborative study. B.P.B. This research was supported in part by the Intramural Research Program of the National Institutes of Health (NIH), the National Eye Institute (EY000564). The contributions of the NIH author(s) were made as part of their official duties as NIH federal employees, are in compliance with agency policy requirements, and are considered Works of the United States Government. However, the findings and conclusions presented in this paper are those of the author(s) and do not necessarily reflect the views of the NIH or the U.S. Department of Health and Human Services. Z.B. NeuroDev study is supported by the Stanley Centre for Psychiatric Research at the Broad Institute and the National Institute of Mental Health (599648 and U01MH119689), National Human Genome Research Institute, the National Eye Institute, and the National Heart, Lung and Blood Institute grant UM1 HG008900 and in part by National Human Genome Research Institute grant R01 HG009141. The content is solely the responsibility of the authors and does not necessarily represent the official views of the National Institutes of Health. D.G.C. This study was supported in part by the US National Human Genome Research Institute (NHGRI) grant as part of the GREGoR Consortium (U01 HG011758 to J.E.P., J.R.L., and R.A.G.) and US National Institute of Neurological Disorders and Stroke (R35NS105078 to J.R.L.) J.X.C. The University of Washington Center for Rare Disease Research (UW-CRDR) is supported by NHGRI grants U01 HG011744 and U24 HG011746. K.A.D. NeuroDev study is supported by the Stanley Center for Psychiatric Research at the Broad Institute, a grant from SFARI (599648, E.B.R), and the National Institute of Mental Health (U01MH119689) and in part by National Human Genome Research Institute grant R01 HG009141 and National Human Genome Research Institute, the National Eye Institute, and the National Heart, Lung and Blood Institute grant UM1 HG008900. The content is solely the responsibility of the authors and does not necessarily represent the official views of the National Institutes of Health L.G. UDP-Vic acknowledges financial support from the Murdoch Children’s Research Institute and the Harbig Foundation. The research conducted at the Murdoch Children’s Research Institute was supported by the Victorian Government’s Operational Infrastructure Support Program R.A.G. B.G. This research was supported in part by the Intramural Research Program of the National Institutes of Health (NIH), the National Eye Institute (EY000564). The contributions of the NIH author(s) were made as part of their official duties as NIH federal employees, are in compliance with agency policy requirements, and are considered Works of the United States Government. However, the findings and conclusions presented in this paper are those of the author(s) and do not necessarily reflect the views of the NIH or the U.S. Department of Health and Human Services. P.K. NeuroDev is supported by the Stanley Center for Psychiatric Research at the Broad Institute. NeuroDev was also supported by a grant from the Simons Foundation or the Simons Foundation International (599648, E.B.R). Research reported in this publication was supported by the: National Institute Of Mental Health of the National Institutes of Health under Award Number U01MH119689; Eunice Kennedy Shriver National Institute Of Child Health & Human Development of the National Institutes of Health under Award Number R01HD102975; National Human Genome Research Institute of the National Institutes of Health under Award Number R01HG012781. The content is solely the responsibility of the authors and does not necessarily represent the official views of the National Institutes of Health. Sequencing was provided by the Broad Institute of MIT and Harvard Center for Mendelian Genomics (Broad CMG) and was funded by the National Human Genome Research Institute; the National Eye Institute; the National Heart, Lung, and Blood Institute grant UM1HG008900; and in part by National Human Genome Research Institute grant R01HG009141. J.R.L. A.S.M. GeneAdd was funded by Luminesce Alliance R.M. RM supported in part by the Samuel Fang Foundation. C.R.N. NeuroDev is supported by the Stanley Center for Psychiatric Research at the Broad Institute. NeuroDev was also supported by a grant from the Simons Foundation or the Simons Foundation International (599648, E.B.R). Research reported in this publication was supported by the: National Institute Of Mental Health of the National Institutes of Health under Award Number U01MH119689; Eunice Kennedy Shriver National Institute Of Child Health & Human Development of the National Institutes of Health under Award Number R01HD102975; National Human Genome Research Institute of the National Institutes of Health under Award Number R01HG012781. The content is solely the responsibility of the authors and does not necessarily represent the official views of the National Institutes of Health. Sequencing was provided by the Broad Institute of MIT and Harvard Center for Mendelian Genomics (Broad CMG) and was funded by the National Human Genome Research Institute; the National Eye Institute; the National Heart, Lung, and Blood Institute grant UM1HG008900; and in part by National Human Genome Research Institute grant R01HG009141. K.Õ. is supported by the Estonian Research Council Grants PRG471 and PRG2040. J.E.P. J.R.L. D.G.C. and R.A.G. were supported in part by the US National Human Genome Research Institute (NHGRI) grant as part of the GREGoR Consortium (U01 HG011758). J.R.L. was supported by the US National Institute of Neurological Disorders and Stroke (R35NS105078) A.R. acknowledges funding from the European Regional Development Fund (ERDF; project NME-GPS) as well as from the “Stiftung Universitätsmedizin Essen” (Duisburg-Essen University). T.R. Australian NHMCR Centre for Research Excellence in Neurocognition 1117394 T.Y.T. UDP-Vic acknowledges financial support from the Murdoch Children’s Research Institute and the Harbig Foundation. The research conducted at the Murdoch Children’s Research Institute was supported by the Victorian Government’s Operational Infrastructure Support Program E.U. This research was supported in part by the Intramural Research Program of the National Institutes of Health (NIH), the National Eye Institute (EY000564). The contributions of the NIH author(s) were made as part of their official duties as NIH federal employees, are in compliance with agency policy requirements, and are considered Works of the United States Government. However, the findings and conclusions presented in this paper are those of the author(s) and do not necessarily reflect the views of the NIH or the U.S. Department of Health and Human Services. C.A.W. was supported by R01NS035129 (NIH/NINDS) and is an Investigator of the Howard Hughes Medical Institute. E.C.E is an Investigator of the Howard Hughes Medical Institute. M.T.W. is supported in part by the National Human Genome Research Institute of the National Institutes of Health through U01HG011762 as part of the GREGoR Consortium. The content is solely the responsibility of the author and does not necessarily represent the official views of the National Institutes of Health. S.M.W. UDP-Vic acknowledges financial support from the Murdoch Children’s Research Institute and the Harbig Foundation. The research conducted at the Murdoch Children’s Research Institute was supported by the Victorian Government’s Operational Infrastructure Support Program. K.M.B. was supported by the National Eye Institute (R01EY035717), P30EY014104 (MEE core support), Foundation Fighting Blindness (BR-GE-0725-0917-MEE). E.A.P is supported by the National Eye Institute (R01EY012910).

## Broad CMG and GREGoR consortium collaborators conflict of interest

A.H.B. has received in-kind research support from Oxford Nanopore Technologies, Pacific Biosciences and GeneDx and is a consultant to Astellas Pharma. S.I.B. is an employee of Ambry Genetics, a subsidiary of Tempus AI.

---

**Supplementary Figure 1:**
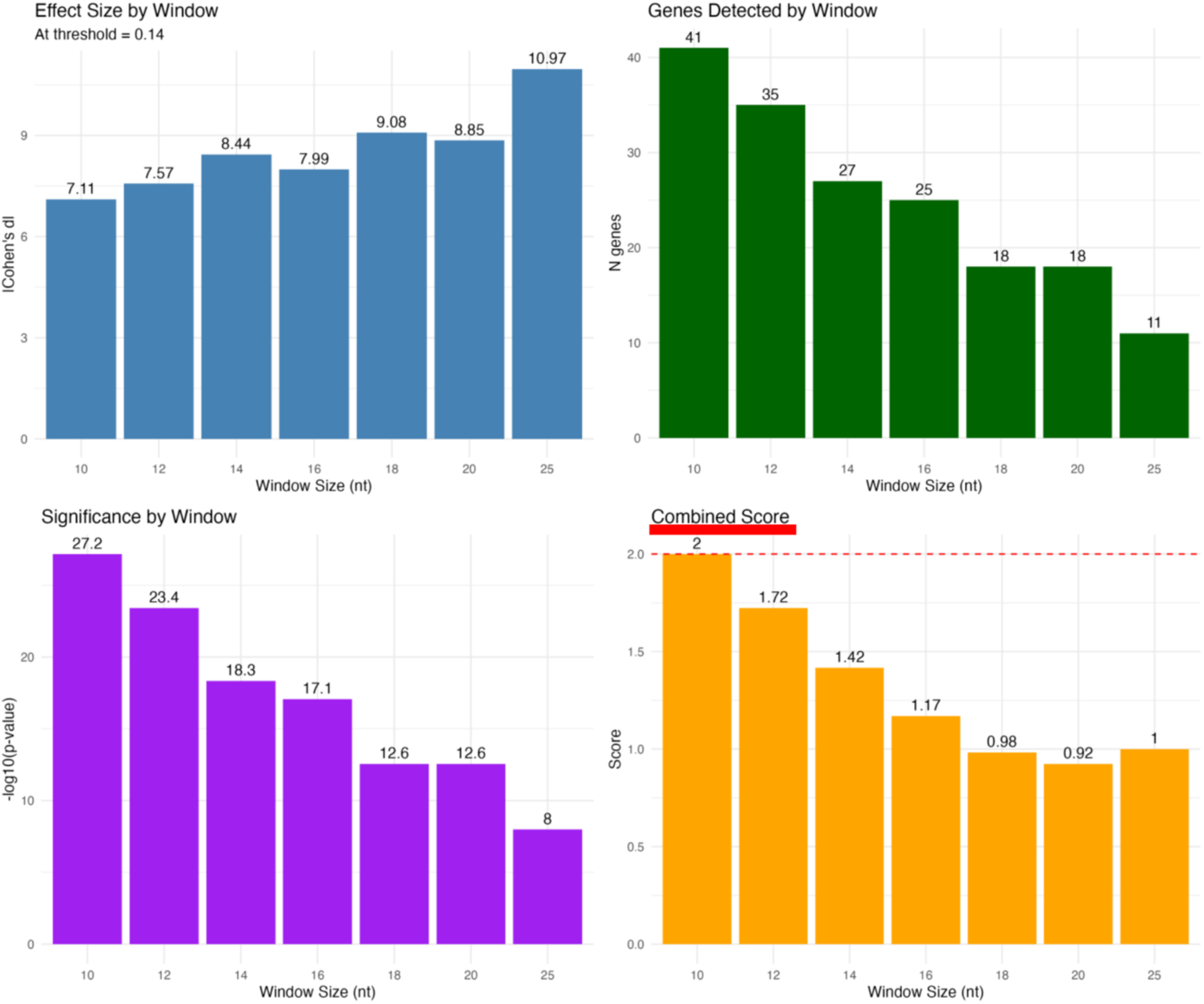
Results of the statistical analysis at threshold 0.14. The number of genes detected (top right), the effect size (Cohen’s d) (top left), the statistical significance (-log10 p-value) (bottom left), and the combined score (bottom right) by window size.

**Supplementary Figure 2:**
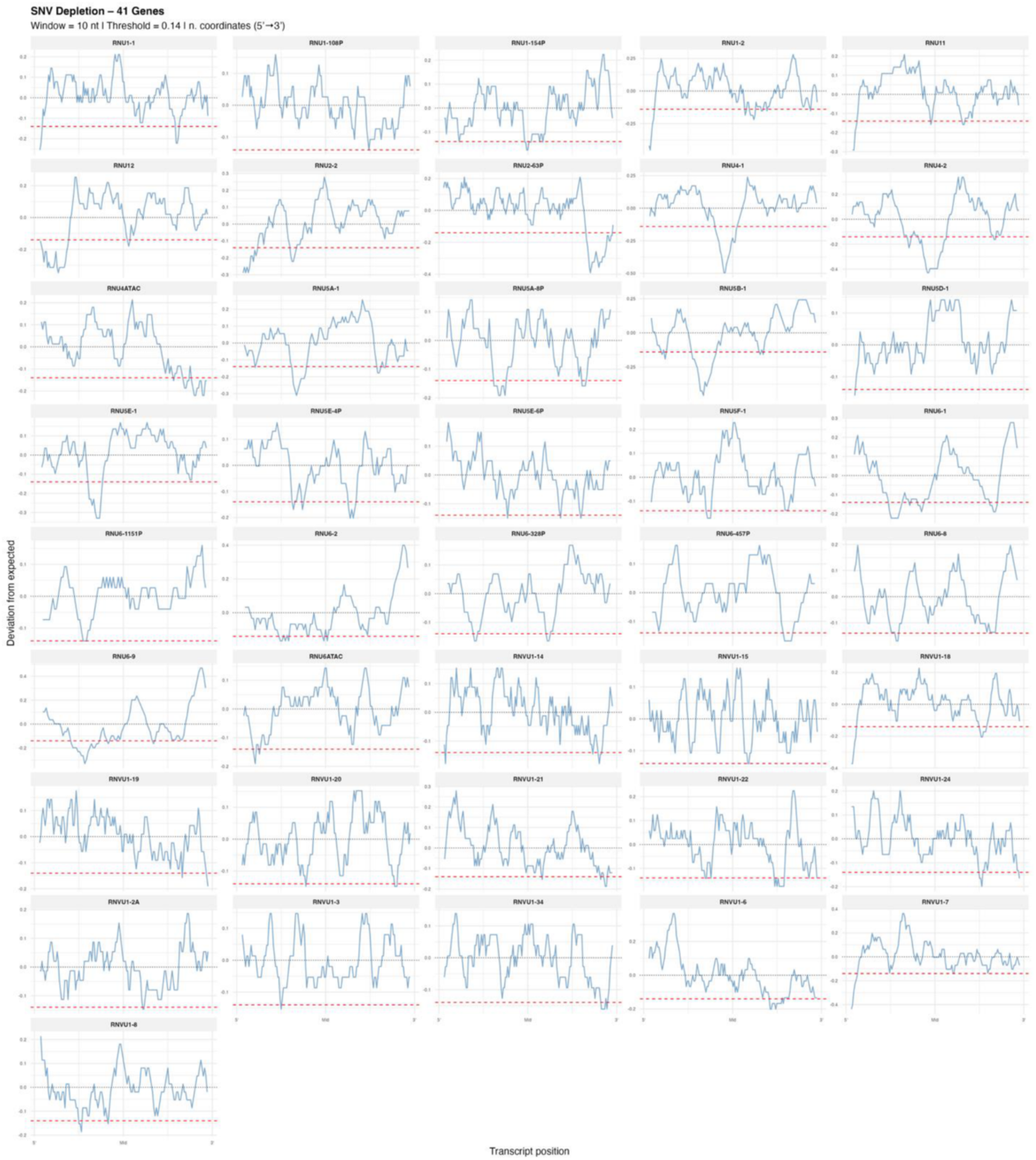
Visualization of depleted regions for all prioritized genes. Plots show the distance to expected by normalized position. The dashed red line indicates the optimal threshold at 0.14.

**Supplementary Figure 3:**
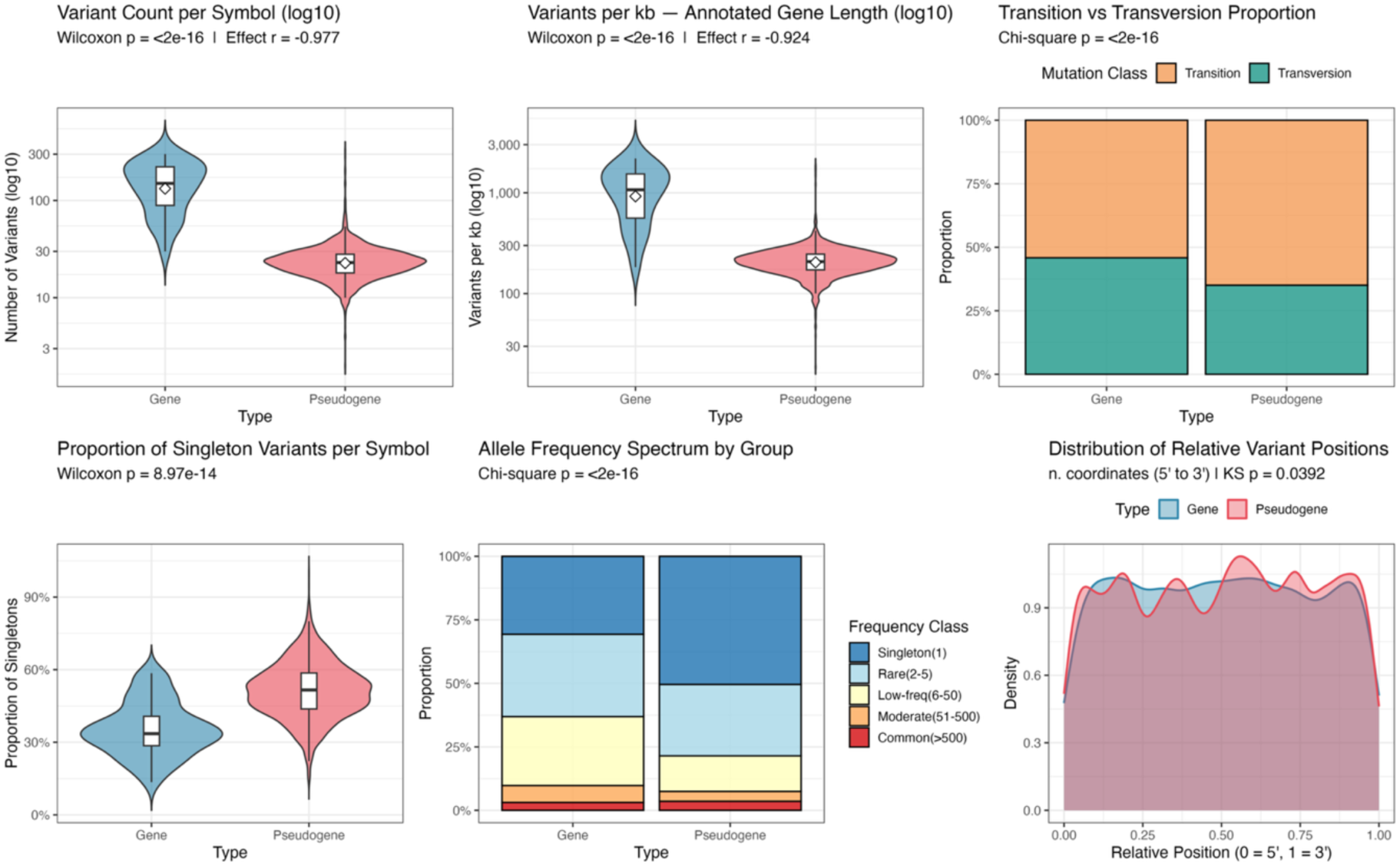
Comparison between snRNA genes and pseudogenes. Plots show that snRNA genes have significantly higher number of variants than pseudogenes per gene (top left) and per kb (top middle), a significant difference in transition vs transversion proportion (top right), singleton proportion (bottom left), and allele frequency (bottom middle). Variant position distribution showed a significant difference in the distribution uniformity (bottom right).

**Supplementary Figure 4:**
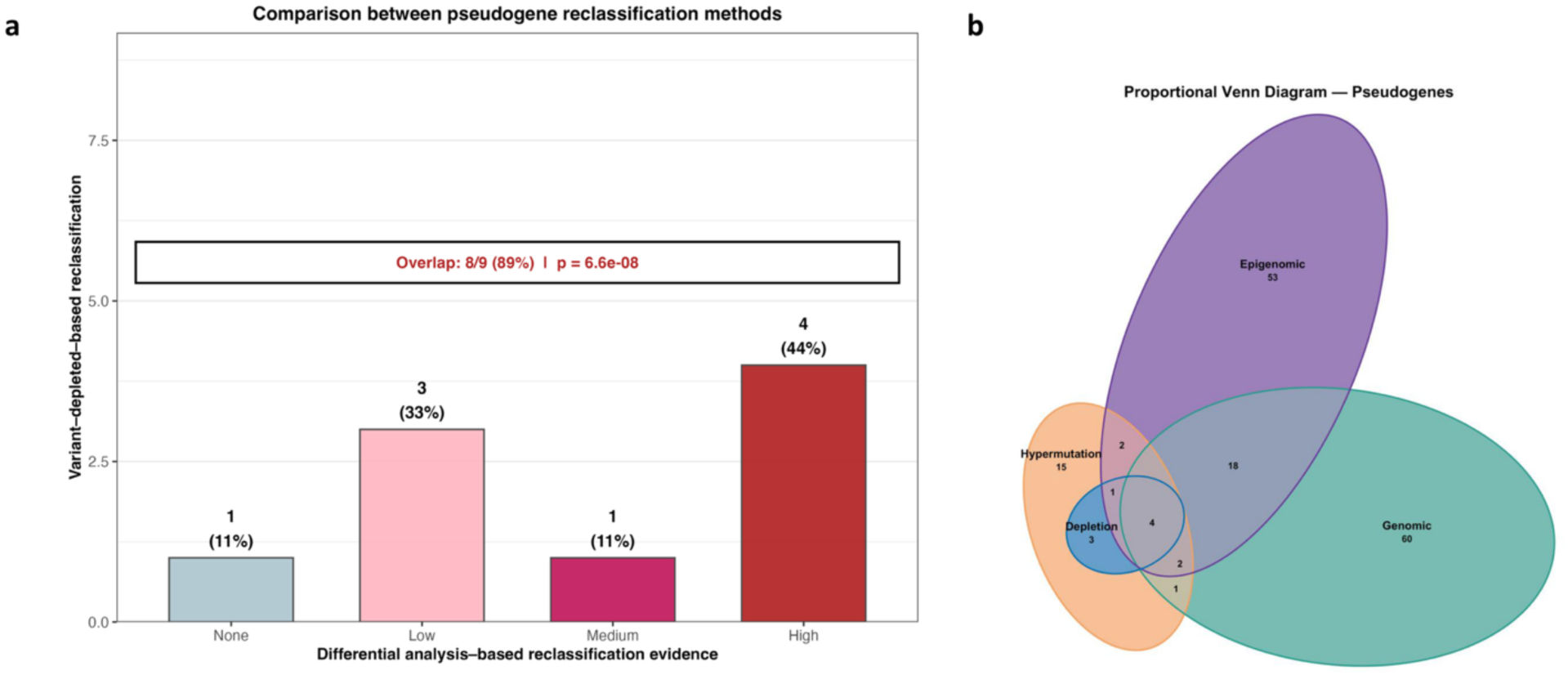
a) **comparison between the variant-depletion approach and the differential approach.** b) **Proportional Venn diagram**. The numbers indicate the gene-like results by the different features of the differential analysis and the depletion analysis.

**Supplementary Figure 5:**
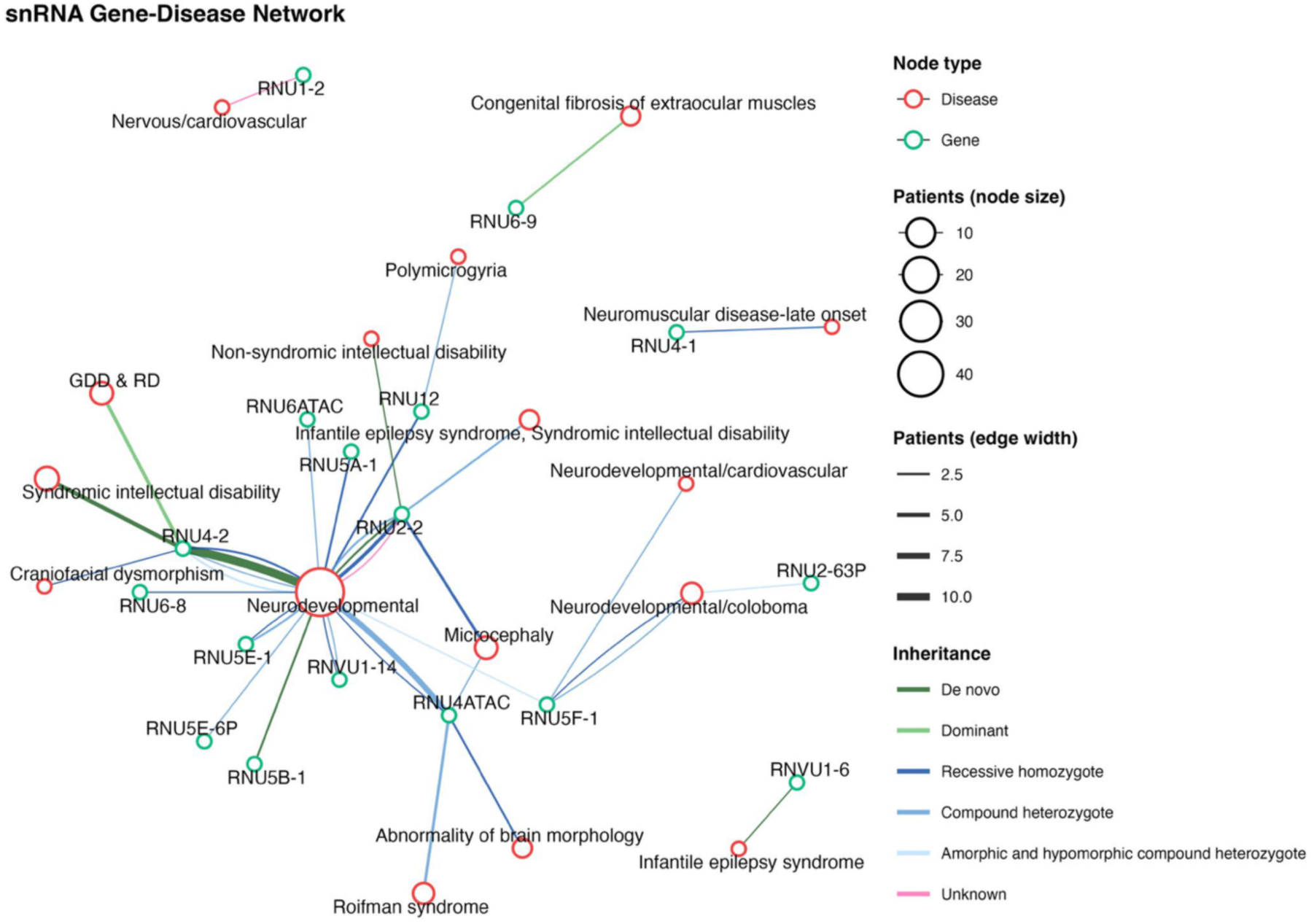
Gene-disease network: The graph shows phenotypic heterogeneity and inheritance variability related to RNU genes and pseudogenes. Green and red circles represent genes and diseases, respectively with the size of the latter being proportional to the number of patients having the corresponding phenotype. The width of the lines is proportional to the number of patients in this gene-phenotype inheritance pattern.

**Supplementary Figure 6:**
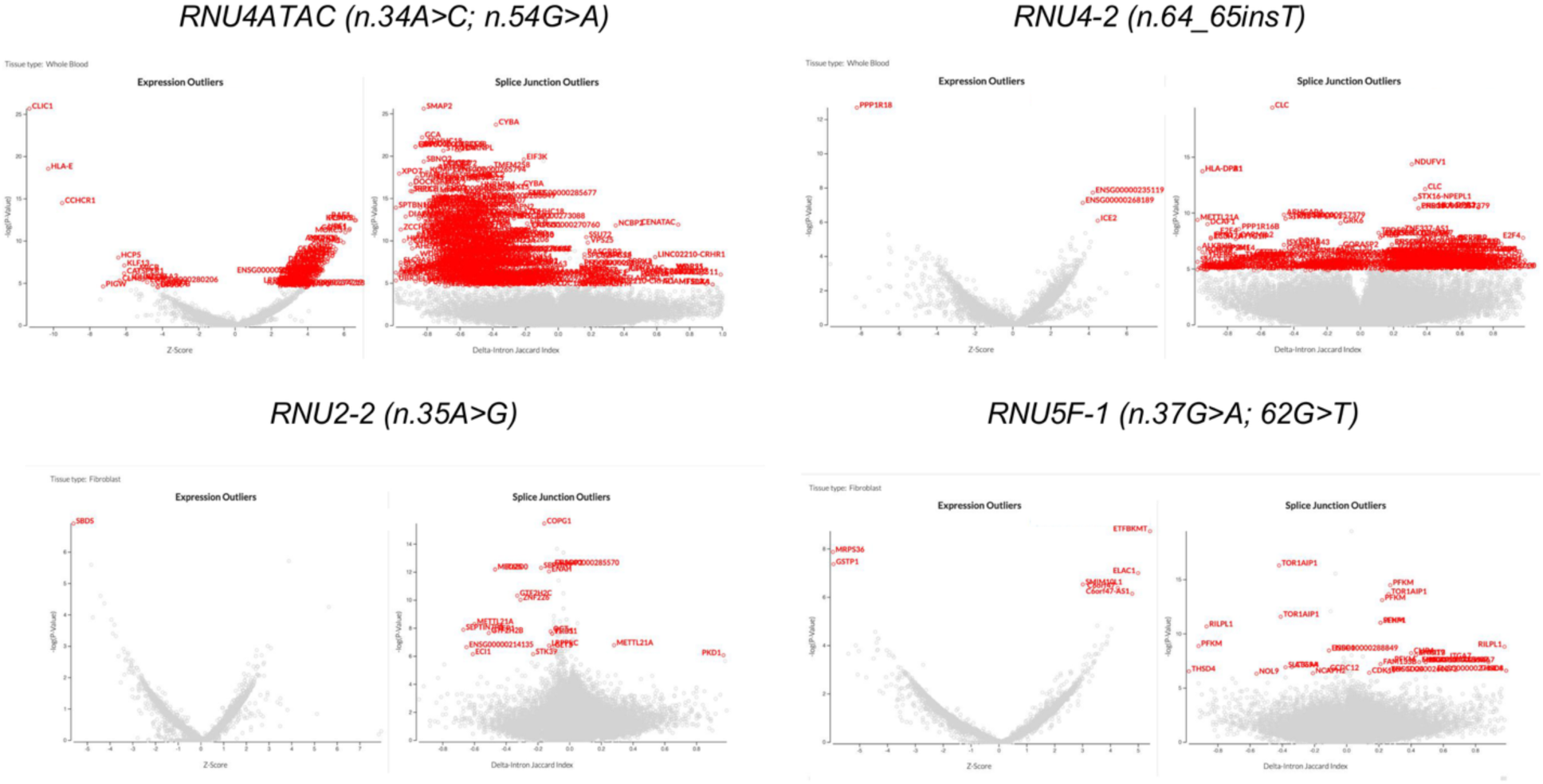
RNAseq data analysis: For each panel, volcano plots illustrate OUTRIDER (left) and FRASER2 (right) results. Genes in red correspond to expression and splice junction outliers (SJO), respectively. Diagnostic variants show significant high SJO in *RNU4ATAC* and *RNU4-2*, but not for *RNU2-2*. Results pertaining to a candidate variant in *RNU5F-1* are also shown.

